# Effect of Enhanced Nutrition and Infection Management Intervention Packages on Antenatal Quality of Care Indicators in Rural Amhara, Ethiopia

**DOI:** 10.64898/2026.07.27.26359070

**Authors:** Nebiyou Fasil, Firehiwot Workneh, Kalkidan Yibeltal, Unmesha Roy Paladhi, Yunhee Kang, Workagegnhu Tarekegn Kidane, Yoseph Yemane Berhane, Fred Van Dyk, Krysten North, Rose L. Molina, Blair J. Wylie, Luke C Mullany, Alemayehu Worku, Anne CC Lee, Yemane Berhane

## Abstract

**Background:** High-quality antenatal care (ANC) can improve the detection, management, and monitoring of pregnancy-related complications. International guidelines recommend at least 8 antenatal care contacts during pregnancy. Interventions to improve the quality of antenatal care may promote positive pregnancy outcomes.

**Objectives:** To assess the impact of enhanced nutrition and infection intervention packages on antenatal quality of care indicators among pregnant women in rural Amhara, Ethiopia

**Methods:** Pregnant women presenting at 12 rural health centers at <24 weeks of gestation were enrolled in this pragmatic clinical effectiveness study. Using 2×2 factorial design, health facilities were allocated to provide an Enhanced Nutrition Package (ENP) or routine nutrition care (non-ENP), followed by individual-level randomization into Enhanced Infection Management Package (EIMP) or routine care (non-EIMP). The composite antenatal quality of care (QoC) score was calculated and compared between arms using cluster-level analyses, at the health center level (for ENP marginal effects), multivariate regression analyses (for EIMP marginal effects), and generalized estimating equations (for combined effects versus routine care). All models were adjusted for imbalanced individual and household factors.

**Results:** From August 2020 to December 2021, a total of 2392 women were randomized (604 ENP+EIMP, 600 ENP alone, 593 EIMP alone, 595 neither package) and followed until June 2022. There was a significant difference in the number of ANC contacts in the ENP group (vs non-ENP: adjusted mean difference [aMD]=1.67, 95% CI: 1.18 to 2.16), EIMP (vs non-EIMP: aMD=0.38, 95% CI: 0.14 to 0.62), and ENP+EIMP (vs routine: aMD=1.92, 95% CI: 1.48 to 2.36). There was also a significant difference in ANC quality of care (ANC QoC) score in ENP (vs non-ENP: aMD=2.13, 95% CI: 0.54 to 3.72); EIMP (vs non-EIMP: aMD=0.89, 95% CI, 0.44 to 1.34), and ENP+EIMP (vs routine: aMD=2.77, 95% CI: 1.28 to 4.27).

**Conclusions:** Combination of enhanced nutrition and infection management interventions had the largest effect on the number of ANC contacts and ANC QoC score. Bolstering nutrition and infection management services during pregnancy could encourage additional ANC contacts, providing an opportunity to provide targeted care.

## Introduction

The World Health Organization (WHO) envisions that every pregnant woman and newborn receive high-quality care throughout pregnancy, childbirth, and the postnatal period worldwide(1,2). Antenatal care (ANC) should encompass four key components: risk identification; prevention and management of pregnancy-related or concurrent diseases; health education and health promotion; and birth preparedness and complication readiness planning, which aim to address health risks, including anemia, malnutrition and infectious diseases(2). These key infection prevention and management, nutritional education and management, and ANC indicators are crucial in assessing the effectiveness of maternal and child health programs in a country.

Maternal nutrition interventions, early identification of malnutrition, and proper treatment of maternal infections improve outcomes for both the mother and the infant (3–5). Studies in low-income settings have shown that high-quality ANC services significantly lower odds of adverse pregnancy outcomes, such as obstructed labor, hypertensive disorders, stillbirth, low-birth weight and preterm births (6–13). High-quality ANC services also promote higher odds of skilled birth attendance and institutional delivery (14). Lifetime risk of adverse birth outcomes is a major public health problem in low-income settings like Ethiopia(15), where maternal mortality and neonatal mortality continue to be high (16–18). A Cochrane review of interventions for improving ANC coverage and health outcomes, including community-based interventions (media campaigns, financial incentives) or health systems interventions (home visits or equipping clinics), showed that there were marginal improvements in ANC coverage with a single package of (interventions aimed at the health system or on target population) interventions compared to no intervention. However, there was limited evidence on the effect of combined interventions on improving ANC quality (19).

In 2016, the WHO recommended eight ANC visits to improve maternal and neonatal outcomes (20), guidance that was subsequently adopted by the Ethiopian Ministry of Health in 2022 (21). Evidence from Ethiopia and other sub-Saharan countries showed that completeness of ANC is vital to addressing pregnancy risks, which would be missed if there were to be fewer contacts (22). Despite improvements in ANC access, there is growing evidence highlighting the need for improved assessment of the quality of ANC services (23), and gaps that exist in adherence to quality services which hamper desired outcomes(22). Successful implementation of recommended maternal nutrition services into ANC services and into the health system (24,25) is one strategy that may impact frequency of contacts and overall ANC quality of care(26).

The Enhancing Nutrition and Antenatal Infection Treatment (ENAT) study aimed to assess the impact of packages of WHO-recommended ANC interventions to optimize maternal nutrition and detect and manage maternal pregnancy infections on infant birth size in Amhara, Ethiopia (27). The aim of this secondary analysis was to determine the effect of maternal nutrition and/or maternal infection management intervention packages on the quality of ANC among pregnant women in rural Amhara, Ethiopia.

## Methods

### Study design

The Enhancing Nutrition and Antenatal Infection Treatment (ENAT) study (ISRCTN15116516) was a 2×2 factorial, pragmatic, open-label randomized clinical effectiveness study. Twelve health centers were cluster-randomized to the Enhanced Nutrition Package (ENP) or the routine nutrition care (non-ENP) arm. All participants were individually randomized to the Enhanced Infection Management Package (EIMP) or the routine infection care (non-EIMP) arm. Therefore, individuals were randomized to one of the four study arms: ENP+EIMP, ENP only, EIMP only, or routine care. The full ENAT study protocol has been published elsewhere(27).

### Participants

The study was conducted in twelve health centers in West Gojam and South Gondar, Amhara, Northwestern Ethiopia, from August 5, 2020 to June 30, 2022. Pregnant women at <24 weeks of gestation visiting ANC clinics who lived within two hours walking distance from the health centers and had a viable fetus on an enrolment ultrasound were included in the study.

### Study Interventions

The enhanced nutrition package (ENP) and enhanced infection management package (EIMP) interventions were delivered through the existing health system. ENP included nutritional counselling, a regular monthly supply of adequately iodized salt, provision of iron-folic acid counseling and supply, and, for undernourished women (mid-upper arm circumference <23 cm), a daily Balanced Energy Protein (BEP) supplement. EIMP included screening and treatment for chlamydia/gonorrhea, bacterial vaginosis and Trichomonas, and deworming with mebendazole at second and third trimester. Women were reminded to attend their scheduled antenatal visits and were also visited at home when they missed their visits (28).

### Outcome Measurement

The quality of antenatal care was assessed using a calculated score (Antenatal Care Quality of Care (ANC QoC), based on previous work done by Countdown in selecting and measuring components of ANC quality in low– and middle-income countries (29–32). Based on the available relevant indicators, the following ten components of care provided at any ANC contact were summed to create the score, with each component scored, no=0 or yes=1, unless specified otherwise. The components were number of ANC contacts, blood pressure measured (at least two times), infection screening (blood sample collection for HIV, syphilis, gonorrhea, chlamydia testing and stool sample collection for parasitic infection testing) done, urine sample collected, tetanus toxoid vaccine (at least two shots) received, deworming received, iron and folic acid received, nutritional counseling received, hemoglobin measurement, completed and counseling on pregnancy danger signs received (Annex-Table 1a). Cronbach’s alpha scale of reliability for the outcome measurement was 0.576, indicating sufficient reliability (33,34).

**Table 1.** Participant, household, and facility characteristics by study arms.

|  | Nutrition allocation |  | Infection allocation |  |
| --- | --- | --- | --- | --- |
| Characteristics | ENP<br>(n=1204) | non-ENP<br>(n=1188) | EIMP<br>(n=1197) | non-EIMP<br>(n=1184) |
| Maternal age, Median (IQR) | 25 (8) | 26 (8) | 25 (8) | 26 (8) |
| Maternal educational status |  |  |  |  |
| No formal education | 590 (49.5) | 563 (47.7) | 577 (48.7) | 576 (48.6) |
| Primary or lower | 357 (30) | 314 (26.6) | 340 (28.7) | 311 (27.9) |
| Secondary or higher | 244 (20.5) | 303 (25.68) | 268 (22.6) | 279 (23.5) |
| Maternal occupational status |  |  |  |  |
| Informal | 292 (24.5) | 378 (32) | 339 (28.6) | 331 (27.9) |
| Agriculture/daily labor | 673 (56.5) | 531 (45) | 604 (50.9) | 600 (50.6) |
| Waged occupation | 227 (19) | 271 (22.9) | 243 (20.5) | 255 (21.5) |
| Maternal GA at study enrolment, Mean (±SD) | 16.4<br>(±5.58) | 16.7 (±5.68) | 16.2<br>(±5.66) | 16.4<br>(±5.59) |
| Maternal parity |  |  |  |  |
| Null (0) | 415 (34.5) | 397 (33.5) | 425 (35.6) | 387 (32.5) |
| Primiparous (1 baby) | 263 (21.9) | 254 (21.4) | 245 (20.5) | 272 (22.8) |
| Multi (2-4 babies) | 368 (30.6) | 388 (32.8) | 384 (32.2) | 372 (31.2) |
| Grand (≥ 5 babies) | 155 (12.91) | 144 (12.1) | 139 (11.6) | 160 (13.4) |
| Current Pregnancy was desired, n(%) | 984 (82.7) | 955 (80.9) | 981 (82.8) | 958 (80.8) |
| Partner educational status |  |  |  |  |
| No formal education | 594 (49.9) | 569 (48.3) | 576 (48.7) | 587(49.6) |
| Primary or lower | 357 (30) | 314 (26.7) | 344 (29.1) | 327 (27.6) |
| Secondary or higher | 239 (20.1) | 294 (25) | 263 (22.2) | 270 (22.8) |
| Land Ownership, n (%) | 812 (68.2) | 848 (71.9) | 834 (70.4) | 826 (69.7) |
| Travel time to health facility (Minutes), Mean (±SD) | 39.4<br>(±60.8) | 43.3<br>(±33.7) | 42.2<br>(±61.9) | 40.5<br>(±32) |

Abbreviations: ENP= enhanced nutrition package, IQR= interquartile range, SD= standard deviation, GA= gestational age.

### Data management

Data were collected by study nurses and data collectors at the health center or at home, using tablets and the Survey Solutions platform (World Bank, V.20.08, 2021). Study staff were trained in data collection protocols, and a field coordinator monitored and supervised study activities on-site. Field staff routinely extracted data from paper-based medical records (Integrated ANC Card & Patient Chart/Card), and the study team routinely conducted data quality control checks.

### Statistical Analysis

In this study, we determined the impact of the ENP and EIMP interventions by comparing marginal effects of ENP vs. non-ENP, EIMP vs. non-EIMP on the number of ANC contacts and ANC QoC score. We also compared the ENP+EIMP vs. routine care (receiving neither package) arms. Cluster-level analysis was used to evaluate the marginal effect of the ENP intervention on the number of ANC contacts and ANC QoC score, and we estimated the mean difference (MD) between ENP and non-ENP groups using t-tests. For dichotomous outcomes, we calculated cluster event rates with log-binomial regression and estimated the relative risk (RR). Multivariable linear regression analysis was used to estimate the marginal effect of EIMP interventions on the number of ANC contacts and ANC QoC score. We also used generalized estimating equations (GEE) to assess the marginal effect of combined ENP+EIMP interventions on the number of ANC contacts and ANC QoC score. All models were adjusted for maternal age, maternal education, maternal occupation, gestational age at enrolment, parity, current pregnancy intention status, and land ownership. Data analysis was conducted using STATA/SE 14.0 (StataCorp, TX, USA) statistical software.

### Ethics statement

This study was approved by Addis Continental Institute of Public Health (001-A1-2019) and Mass General Brigham Institutional Review Board (2018P002479). Participants provided informed written consent to participate in the study before completing any study procedures.

## RESULTS

### Participant characteristics

Among 3145 pregnant women screened between August 2020 and December 2021, 2392 were eligible and randomized into the four study arms (Figure 1). Several characteristics, including maternal education, maternal occupation, and land ownership, were imbalanced between the ENP and non-ENP arms (Table 1). Participant characteristics were similar, however, across EIMP and non-EIMP arms. The combined intervention group arm (ENP+EIMP) had an imbalance in participants’ maternal education, maternal occupation, and land ownership characteristics (Annex-Table 1b).

**Figure 1.**
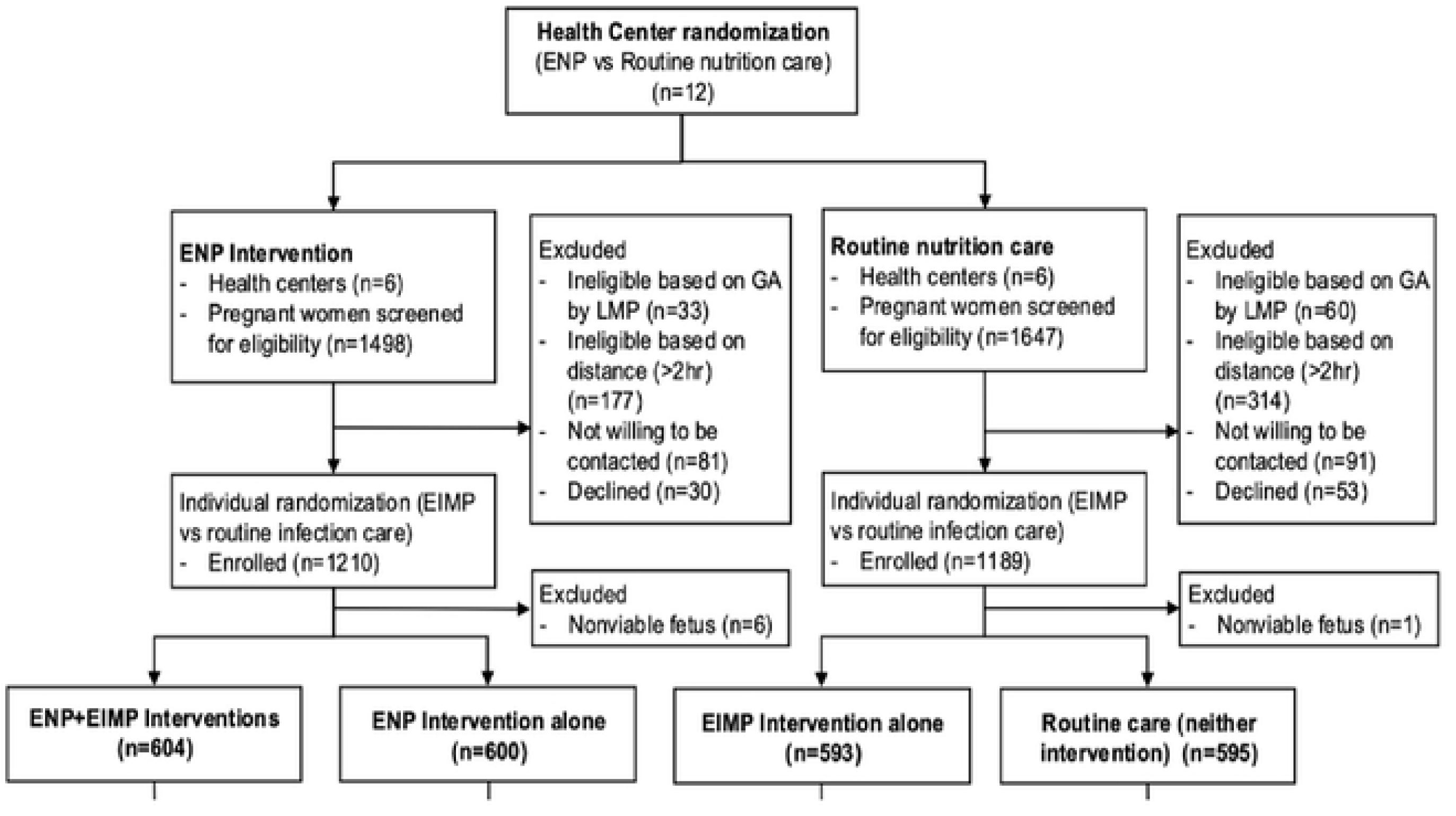
CONSORT diagram of participants in the ENAT study.

### Number of ANC contacts and quality of care indicators

The ENP arm had a higher number of ANC contacts (mean=4.3, standard deviation (SD)=1.7) compared to the non-ENP arm (mean=2.8, SD=1.2) (adjusted mean difference (aMD)=1.63, 95% CI: 1.13 to 2.13) (Table 2). Blood pressure measurement frequency was significantly higher in the ENP arm compared to the non-ENP arm. Women in the ENP arm had a significantly higher ANC QoC score (aMD=2.13, 95% CI: 0.54 to 3.72).

**Table 2.**
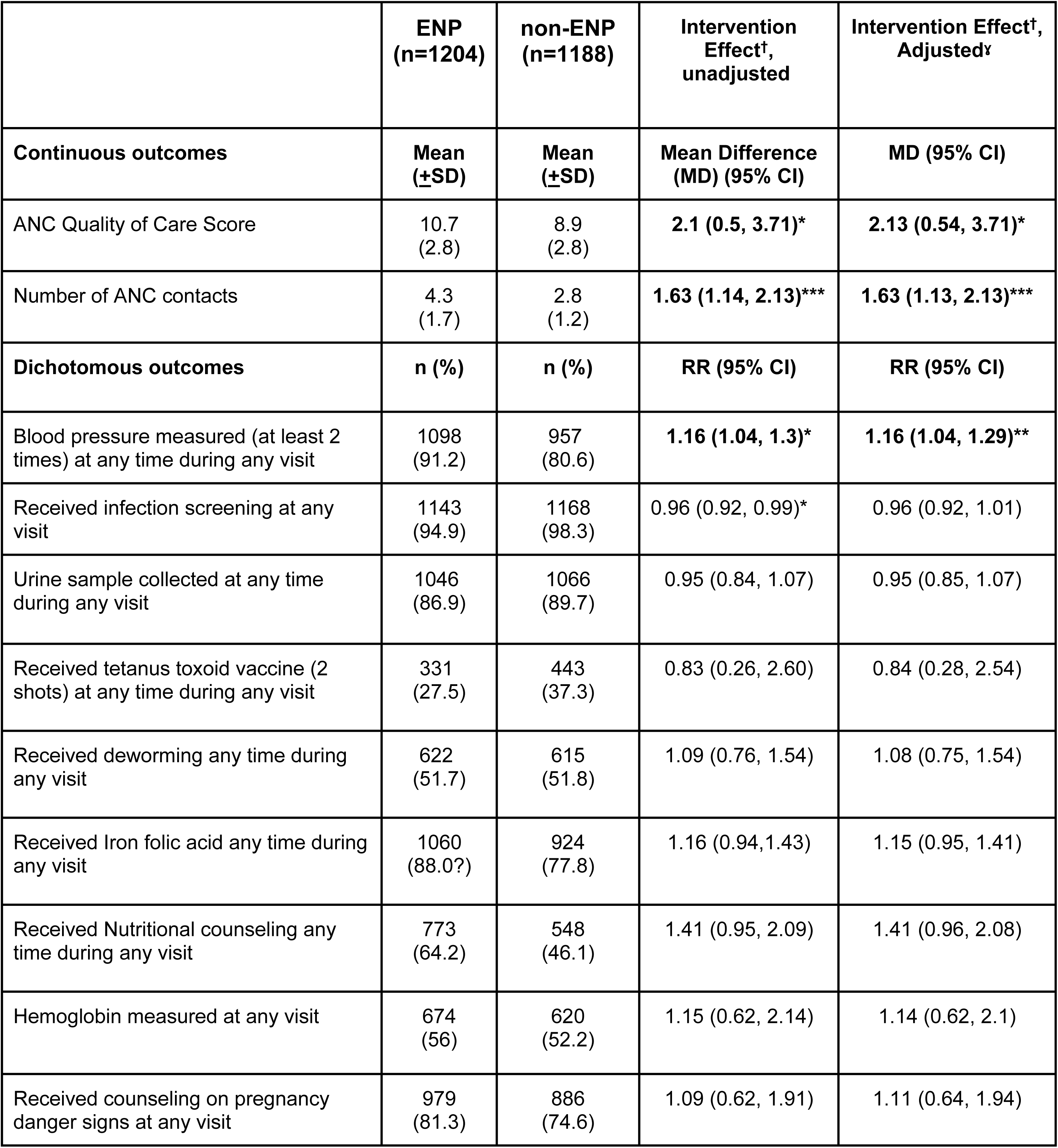
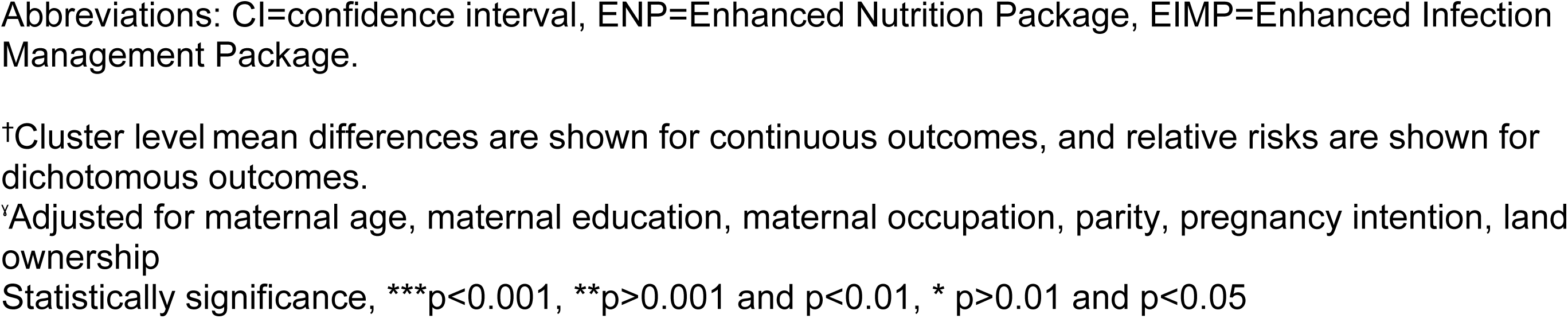
Effects of Enhanced Nutrition Package on ANC quality of care indicators.

|  | **ENP**<br>**(n=1204)** | **non-ENP**<br>**(n=1188)** | **Intervention**<br>**Effect<sup>†</sup>,**<br>**unadjusted** | **Intervention Effect<sup>†</sup>,**<br>**Adjusted<sup>\*</sup>** |
| --- | --- | --- | --- | --- |
| **Continuous outcomes** | **Mean**<br>**(±SD)** | **Mean**<br>**(±SD)** | **Mean Difference**<br>**(MD) (95% CI)** | **MD (95% CI)** |
| ANC Quality of Care Score | 10.7<br>(2.8) | 8.9<br>(2.8) | **2.1 (0.5, 3.71)\*** | **2.13 (0.54, 3.71)\*** |
| Number of ANC contacts | 4.3<br>(1.7) | 2.8<br>(1.2) | **1.63 (1.14, 2.13)\*\*\*** | **1.63 (1.13, 2.13)\*\*\*** |
| **Dichotomous outcomes** | **n (%)** | **n (%)** | **RR (95% CI)** | **RR (95% CI)** |
| Blood pressure measured (at least 2 times) at any time during any visit | 1098<br>(91.2) | 957<br>(80.6) | **1.16 (1.04, 1.3)\*** | **1.16 (1.04, 1.29)\*\*** |
| Received infection screening at any visit | 1143<br>(94.9) | 1168<br>(98.3) | 0.96 (0.92, 0.99)* | 0.96 (0.92, 1.01) |
| Urine sample collected at any time during any visit | 1046<br>(86.9) | 1066<br>(89.7) | 0.95 (0.84, 1.07) | 0.95 (0.85, 1.07) |
| Received tetanus toxoid vaccine (2 shots) at any time during any visit | 331<br>(27.5) | 443<br>(37.3) | 0.83 (0.26, 2.60) | 0.84 (0.28, 2.54) |
| Received deworming any time during any visit | 622<br>(51.7) | 615<br>(51.8) | 1.09 (0.76, 1.54) | 1.08 (0.75, 1.54) |
| Received Iron folic acid any time during any visit | 1060<br>(88.0?) | 924<br>(77.8) | 1.16 (0.94, 1.43) | 1.15 (0.95, 1.41) |
| Received Nutritional counseling any time during any visit | 773<br>(64.2) | 548<br>(46.1) | 1.41 (0.95, 2.09) | 1.41 (0.96, 2.08) |
| Hemoglobin measured at any visit | 674<br>(56) | 620<br>(52.2) | 1.15 (0.62, 2.14) | 1.14 (0.62, 2.1) |
| Received counseling on pregnancy danger signs at any visit | 979<br>(81.3) | 886<br>(74.6) | 1.09 (0.62, 1.91) | 1.11 (0.64, 1.94) |

Abbreviations: CI=confidence interval, ENP=Enhanced Nutrition Package, EIMP=Enhanced Infection Management Package.
†Cluster level mean differences are shown for continuous outcomes, and relative risks are shown for dichotomous outcomes.
‡Adjusted for maternal age, maternal education, maternal occupation, parity, pregnancy intention, land ownership
Statistically significance, \*\*\*p<0.001, \*\*p>0.001 and p<0.01, \* p>0.01 and p<0.05

Women in the EIMP arm had a similar number of ANC contacts (mean: 3.8, SD: 1.7), to those in the non-EIMP arm (mean: 3.4, SD: 1.5, aMD=0.38, 95% CI: 0.14 to 0.62) (Table 3). Urine sample collection (aRR=1.1, 95% CI: 1.03 to 1.15) and deworming (aRR=2.61, 95% CI: 1.75 to 3.89) were significantly higher in the EIMP arm compared to the non-EIMP arm. Women in the EIMP arm also had higher ANC QoC score compared to those in the non-EIMP arm (aMD=0.89, 95% CI: 0.44 to 1.34).

**Table 3.** Effects of Enhanced Infection Management Package on ANC quality of care indicators.

|  | **EIMP**<br>**(n=1197)** | **non-**<br>**EIMP**<br>**(n=1184)** | **Intervention Effect†,**<br>**unadjusted** | **Intervention Effect†,**<br>**Adjusted‡** |
| --- | --- | --- | --- | --- |
| **Continuous outcomes** | **Mean**<br>**(±SD)** | **Mean**<br>**(±SD)** | **MD (95% CI)** | **MD (95% CI)** |
| ANC Quality of Care Score | 10.3<br>(3.1) | 9.4<br>(2.8) | **0.89 (0.40, 1.39)\*\*** | **0.89 (0.44, 1.34)\*\*** |
| Number of ANC contacts | 3.8<br>(1.7) | 3.4<br>(1.5) | **0.37 (0.13, 0.62)\*\*** | **0.38 (0.14, 0.62)\*\*** |
| **Dichotomous outcomes** | **n (%)** | **n (%)** | **RR (95% CI)** | **RR (95% CI)** |
| Blood pressure measured (at least 2 times) at any time during any visit | 1017<br>(85) | 1038<br>(86.9) | 0.99 (0.99, 0.99) | 0.98 (0.95, 1.01) |
| Received infection screening at any visit | 1176<br>(98.3) | 1135<br>(94.9) | 1.01 (1.001, 1.02)*** | 1.00 (1.00, 1.00) |
| Urine sample collected at any time during any visit | 1105<br>(92.3) | 1007<br>(84.3) | **1.09 (1.03, 1.16)\*\*** | **1.1 (1.03, 1.15)\*\*** |
| Received tetanus toxoid vaccine (2 shots) at any time during any visit | 390<br>(32.6) | 384<br>(32.1) | 1.01 (0.92, 1.12) | 0.99 (0.91, 1.09) |
| Received deworming any time during any visit | 894<br>(74.7) | 343<br>(28.7) | **2.6 (1.74, 3.89)\*\*\*** | **2.61 (1.75, 3.89)\*\*\*** |
| Received Iron folic acid any time during any visit | 980<br>(81.8) | 1004<br>(84) | 0.97 (0.94, 1.01) | 0.97 (0.95, 0.99) |
| Received Nutritional counseling any time during any visit | 657<br>(54.9) | 664<br>(55.6) | 0.98 (0.91, 1.07) | 0.98 (0.92, 1.06) |
| Hemoglobin measured at any visit | 658 (55) | 636<br>(53.2) | 1.03 (0.98, 1.09) | 1.03 (0.97, 1.09) |
| Received counseling on pregnancy danger signs at any visit | 916<br>(76.5) | 949<br>(79.4) | **0.96 (0.93, 0.99)\*** | **0.96 (0.92, 0.99)\*** |

Abbreviations: CI=confidence interval, ENP=Enhanced Nutrition Package, EIMP=Enhanced Infection Management Package.
†Mean differences are shown for continuous outcomes, and relative risks are shown for dichotomous outcomes.
‡Adjusted for maternal age, maternal education, maternal occupation, parity, pregnancy intention, land ownership
Statistically significant, \*\*\*p<0.001, \*\*p>0.001 and p<0.01, \* p>0.01 and p<0.05

Women in the combined ENP+EIMP arm had a higher number of ANC contacts (mean: 4.6, SD: 1.7), compared to the routine care arm (mean: 2.7, SD:1.1), aMD=1.92, 95% CI: 1.48 to 2.36) (Table 4). Women in the combined ENP+EIMP arm had a significantly higher rate of receiving deworming (aRR=2.77, 95% CI: 1.28 to 4.27) and ANC QoC scores (aMD=2.77, 95% CI: 1.28 to 4.27), compared to those in the routine care arm.

**Table 4.** Effects of the combined Enhanced Nutrition and infection management Package on ANC Quality of Care indicators.

|  | ENP+EIMP<br>(n=604) | Routine<br>care<br>(n=595) | Intervention<br>Effect†,<br>unadjusted | Intervention Effect†,<br>Adjusted‡ |
| --- | --- | --- | --- | --- |
| Continuous outcomes | Mean<br>(±SD) | Mean (±SD) | MD (95% CI) | MD (95% CI) |
| ANC Quality of Care Score | 11.3<br>(2.9) | 8.6<br>(2.7) | **2.74 (1.16, 4.32)\*\*** | **2.77 (1.28, 4.27)\*\*\*** |
| Number of ANC contacts | 4.6<br>(1.7) | 2.7<br>(1.1) | **1.89 (1.46, 2.32)\*\*\*** | **1.92 (1.48, 2.36)\*\*\*** |
| Dichotomous outcomes | n (%) | n (%) | RR (95% CI) | RR (95% CI) |
| Blood pressure measured (at least 2 times) at any time during any visit | 552<br>(91.4) | 492<br>(82.7) | 1.1 (1.01, 1.20)* | 1.00 (0.88, 1.14) |
| Received infection screening at any visit | 591<br>(97.8) | 583<br>(97.8) | 0.99 (0.98, 1.02) | 0.98 (0.97, 1.01) |
| Urine sample collected at any time during any visit | 551<br>(91.2) | 512<br>(86.1) | 1.06 (0.96, 1.17) | 1.04 (0.95, 1.14) |
| Received tetanus toxoid vaccine (2 shots) at any time during any visit | 175<br>(28.9) | 228<br>(38.3) | 0.75 (0.29, 1.92) | 0.74 (0.29, 1.88) |
| Received deworming any time during any visit | 489<br>(80.9) | 210<br>(35.3) | **2.29 (1.25, 4.21)\*\*** | **2.29 (1.26, 4.17)\*\*** |
| Received Iron folic acid any time during any visit | 526<br>(87.1) | 470<br>(79) | 1.1 (0.91, 1.34) | 1.09 (0.94, 1.27) |
| Received Nutritional counseling any time during any visit | 393<br>(65.1) | 284<br>(47.7) | 1.36 (0.94, 1.97) | 1.34 (0.96, 1.89) |
| Hemoglobin measured at any visit | 336<br>(55.6) | 298<br>(50.1) | 1.12 (0.68, 1.83) | 1.11 (0.71, 1.75) |
| Received counseling on pregnancy danger signs at any visit | 480<br>(79.5) | 450<br>(75.6) | 1.05 (0.63, 1.73) | 1.05 (0.68, 1.62) |

CI=confidence interval. ENP=Enhanced Nutrition Package. EIMP=Enhanced Infection Management Package
†Mean differences are shown for continuous outcomes, and relative risks are shown for dichotomous outcomes.
‡Adjusted for maternal age, maternal education, maternal occupation, parity, pregnancy intention, land ownership
Statistically significant, \*\*\*p<0.001, \*\*p>0.001 and p<0.01, \* p>0.01 and p<0.05

## Discussion

Our study investigated the effect of nutrition and/or infection management on the number of ANC contacts and the quality of those contacts, as measured through a composite score.

The enhanced antenatal intervention packages provided within the Ethiopian health system increased coverage of and demand for ANC during challenging times for the health system with COVID-19 and regional conflicts. In the enhanced nutrition package (ENP) arm, there was a significant increase in the number of ANC contacts, as well as the proportion of blood pressure measurement, and the overall ANC QoC score. In the enhanced infection management package (EIMP) arm, we also saw an increase in both metrics as well as urine screening with increased urine sample collection and deworming compared to the routine (non-EIMP) arm. In the combined nutrition and infection arm, we saw the most improvement which increased the number of ANC contacts by two times and quality of care score by 2.7. Overall, our study demonstrates the impact of our interventions on improving the quality and coverage of antenatal health services.

Participants in the ENP arm had significantly more ANC contacts than those in the routine arm (aMD = 1.63, 95% CI: 1.13 to 2.13) and a significantly higher ANC quality-of-care score (aMD = 2.13; 95% CI: 0.54 to3.71). The provision of nutrition interventions, including nutritional counseling, household iodized salt, and balanced energy-protein (BEP) supplements, appeared to increase demand for ANC services, as women were motivated to return regularly to receive monthly supplies of iodized salt and BEP. These repeated visits facilitated closer follow-up and created opportunities for the delivery of comprehensive ANC services, including nutrition counseling, blood pressure measurement, nutrition monitoring, and supplement provision. Previous evidence suggests that nutrition information and malnutrition management interventions improve client satisfaction and increase ANC utilization (35,36). Consistent with our findings, a study in Burkina Faso reported higher ANC attendance among women receiving nutrition interventions, including counseling, iron-folic acid supplementation, and weight monitoring, while studies from Bangladesh have demonstrated improvements in ANC quality indicators such as nutritional counseling, IFA provision, and weight monitoring following the integration of nutrition interventions into ANC services (37). Together, these findings suggest that the provision of nutrition supplements and counseling may be an effective strategy for increasing both the coverage of recommended ANC contacts and the quality of care received during pregnancy(30,38).

Participants in the EIMP arm had significantly more ANC contacts than those in the routine care arm (aMD = 0.38; 95% CI: 0.14 to 0.62) and a significantly higher ANC quality-of-care score (aMD = 0.89; 95% CI: 0.44 to 1.34). The integration of universal infection screening and treatment into ANC may have facilitated greater client engagement, follow-up, and continuity of care, contributing to increased ANC attendance. This finding is consistent with evidence from the 2019 Ethiopian Demographic and Health Survey, which showed that women with more frequent ANC contacts were more likely to receive recommended infection screening and management services (39,40). In addition, uptake of key infection-related services was significantly higher in the EIMP arm, including urine sample collection (aRR = 1.10; 95% CI: 1.03–1.15) and deworming (aRR = 2.61; 95% CI: 1.75–3.89). Similar findings have been reported in comparable settings, were infection management packages improved utilization of urine testing and deworming services among pregnant women. Together, these findings suggest that integrating infection prevention, screening, and treatment interventions within routine ANC can enhance both service uptake and the overall quality of antenatal care (6,41).

Participants in the combined Nutrition–Infection Management (ENP+EIMP) arm had significantly higher ANC contacts compared to those in the routine arm (aMD = 1.92; 95% CI: 1.48 to 2.36) as well as a higher antenatal quality-of-care (ANC QoC) score (aMD = 2.77; 95% CI: 1.28 to4.27). Evidence from low– and middle-income countries indicates that more frequent ANC visits are associated with greater utilization of essential nutrition and infection management services that are critical for high-quality care (42), while in Ethiopia, women with fewer ANC contacts are more likely to miss key ANC components (43). Similarly, intervention studies in rural Ethiopia have demonstrated that integrated nutrition and infection management approaches increase uptake of core ANC services, including blood and urine testing, iron–folate supplementation, tetanus toxoid vaccination, deworming, and counseling on nutrition and pregnancy complications (44). Overall, these findings suggest that comprehensive integration of nutrition counseling, supplementation, and infection screening and management within ANC services can promote more frequent visits and improve the overall quality of antenatal care (44,45).

There were several strengths of our study. The interventions were delivered by health system staff to test the effectiveness of pragmatic, real-world implementation. Participants were also randomized into intervention arms to minimize possible biases. Data were also abstracted from participants’ medical records, so it reflects statistics in the health system. The indicators compiled and selected for measuring ANC QoC were extensive and incorporated both nutrition and infection management indicators that were not incorporated in other previous literature.

Limitations affecting the original study, such as the pandemic and civil conflict, have limited-service delivery, field access and follow-up visits during the study period. Hence, our ability to measure ANC services uptake throughout the pregnancy period was compromised. Moreover, when measuring ANC quality, we were not able to capture waiting time, which is an important component of quality service that can lead to dissatisfaction, consequently, affecting compliance to follow up (46). Furthermore, the scale reliability coefficient of ANC QoC score (alpha=0.576) was sufficient, but further enrichment of the scale is required to construct a robust measurement indicator for ANC quality.

## Conclusion

In rural Amhara, Ethiopia, both enhanced nutrition and infection interventions had a positive impact on improving the number of ANC contacts and antenatal quality of care score. Enhancing nutrition and infection services could help drive care-seeking and attain recommended antenatal care contacts, by improving access to key antenatal quality of care services. These findings also emphasize the need for adapting these quality-of-care components for monitoring and evaluation of antenatal care programs.

## Acknowledgements

We thank all families, study participants, study communities, and our dedicated field team. We would like to acknowledge our partners at the Amhara Regional Health Bureau and Amhara Public Health Institute, the Ethiopian Ministry of Health, the Ethiopian Society of Obstetrics and Gynecology, and General Electric. We also acknowledge Jhpiego and the Children’s Investment Fund for support during the codesign phase of the study. We thank Sophie Driker for assistance with manuscript formatting/preparation for submission. We thank our Study Monitoring Committee, Dr. Delayehu Bekele, and Professor Simon Cousens.

## Author contributions

Conceptualization and design: Anne CC Lee, Yemane Berhane, Alemayehu Worku, Luke C. Mullany, Firehiwot Workneh, Rose L. Molina, Blair J. Wylie

Intervention development and delivery: Firehiwot Workneh, Kalkidan Yibeltal, Workagegnhu Tarekegn Kidane, Alemayehu Worku

Project administration, management, and supervision: Firehiwot Workneh, Kalkidan Yibeltal, Workagegnhu Tarekegn Kidane, Nebiyou Fasil, Yoseph Yemane Berhane

Data acquisition and curation; Firehiwot Workneh, Yunhee Kang, Kalkidan Yibeltal, Yoseph Yemane Berhane, Nebiyou Fasil, Fred Van Dyk, Unmesha Paladhi, Kristen North

Statistical analysis: Nebiyou Fasil, Unmesha Roy Paladhi

Writing – original draft: Nebiyou Fasil

Writing – review & editing: Nebiyou Fasil, Firehiwot Workneh, Kalkidan Yibeltal, Unmesha Roy Paladhi, Yunhee Kang, Workagegnhu Tarekegn Kidane, Yoseph Yemane Berhane, Fred Van Dyk, Krysten North, Rose L. Molina, Blair J. Wylie, Luke C Mullany, Alemayehu Worku, Anne CC Lee, and Yemane Berhane

## Data availability statement

Data described in the manuscript, codebook, and analytic code will be made available upon request, pending application and approval.

## Funding

This work was funded by the Bill and Melinda Gates Foundation (OPP1184363/INV-010109, INV-045894). The funder had no role in study design, data collection and analysis, the decision to publish, or preparation of the manuscript.

## Competing interests

Anne CC Lee received support from the Eunice Kennedy Shriver National Institute of Child Health and Human Development (5K23HD091390), World Health Organization, and Johns Hopkins University. All other authors report no conflicts of Interest.

## Supplemental figures and tables

**Table 1a:**
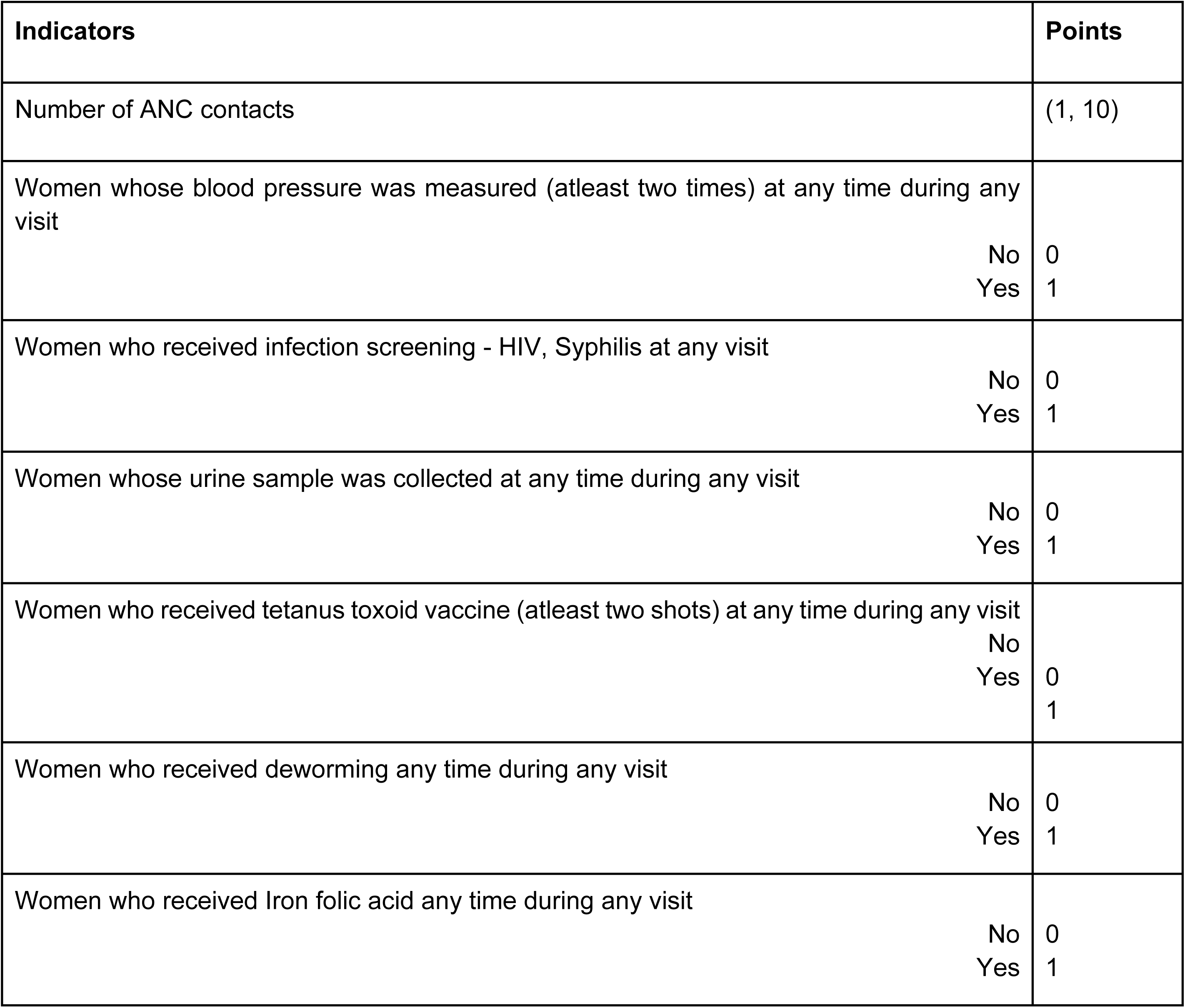
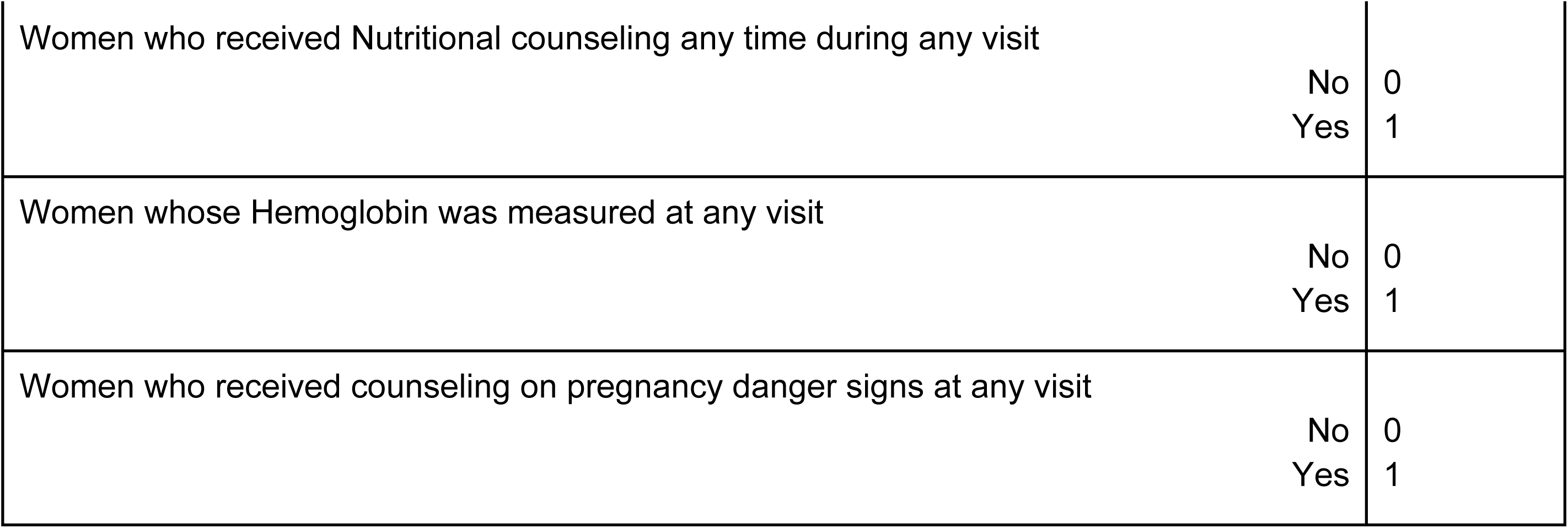
Antenatal Quality of Care (ANC QoC) components scoring.

**Table 1b.**
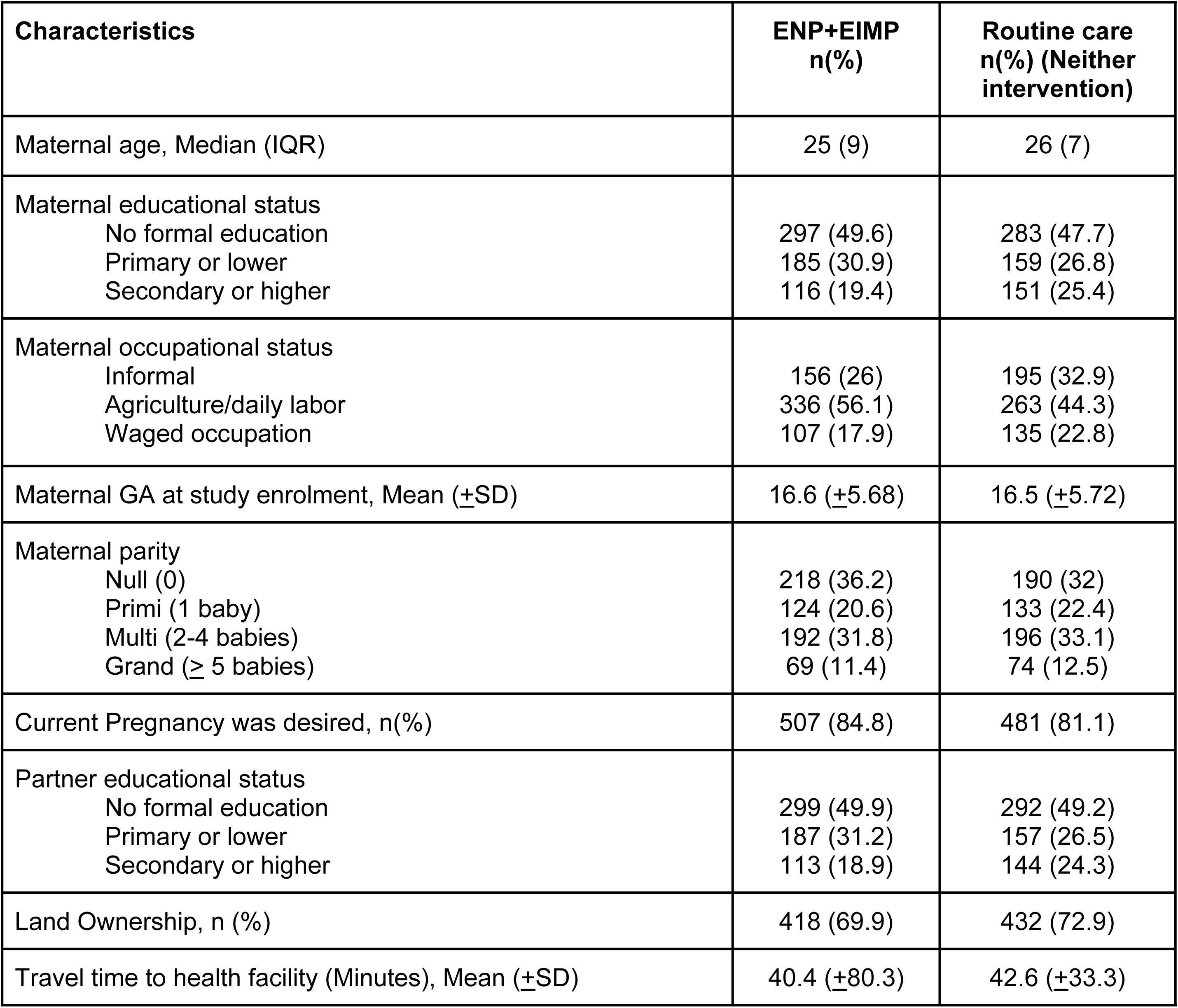
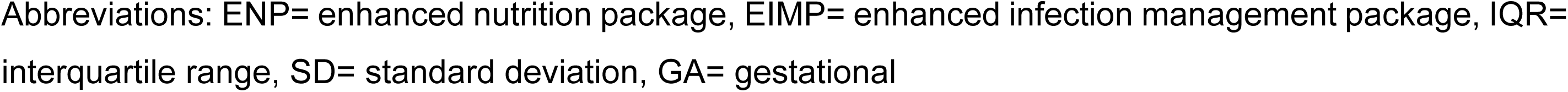
– Participant, household, and facility characteristics for ENP+EIMP vs. routine care arms.

| Characteristics | ENP+EIMP<br>n(%) | Routine care<br>n(%) (Neither<br>intervention) |
| --- | --- | --- |
| Maternal age, Median (IQR) | 25 (9) | 26 (7) |
| Maternal educational status |  |  |
| No formal education | 297 (49.6) | 283 (47.7) |
| Primary or lower | 185 (30.9) | 159 (26.8) |
| Secondary or higher | 116 (19.4) | 151 (25.4) |
| Maternal occupational status |  |  |
| Informal | 156 (26) | 195 (32.9) |
| Agriculture/daily labor | 336 (56.1) | 263 (44.3) |
| Waged occupation | 107 (17.9) | 135 (22.8) |
| Maternal GA at study enrolment, Mean (±SD) | 16.6 (±5.68) | 16.5 (±5.72) |
| Maternal parity |  |  |
| Null (0) | 218 (36.2) | 190 (32) |
| Primi (1 baby) | 124 (20.6) | 133 (22.4) |
| Multi (2-4 babies) | 192 (31.8) | 196 (33.1) |
| Grand (≥ 5 babies) | 69 (11.4) | 74 (12.5) |
| Current Pregnancy was desired, n(%) | 507 (84.8) | 481 (81.1) |
| Partner educational status |  |  |
| No formal education | 299 (49.9) | 292 (49.2) |
| Primary or lower | 187 (31.2) | 157 (26.5) |
| Secondary or higher | 113 (18.9) | 144 (24.3) |
| Land Ownership, n (%) | 418 (69.9) | 432 (72.9) |
| Travel time to health facility (Minutes), Mean (±SD) | 40.4 (±80.3) | 42.6 (±33.3) |

Abbreviations: ENP= enhanced nutrition package, EIMP= enhanced infection management package, IQR= interquartile range, SD= standard deviation, GA= gestational

## Notes

### Clinical Trial

ISRCTN15116516

### Clinical Protocols

https://bmjpaedsopen.bmj.com/content/6/1/e001327

